# Influence of sex, season and environmental air quality on experimental human pneumococcal carriage acquisition

**DOI:** 10.1101/2021.10.04.21264228

**Authors:** Katerina S. Cheliotis, Christopher P. Jewell, Carla Solórzano, Britta Urban, Andrea M. Collins, Elena Mitsi, Sherin Pojar, Elissavet Nikolaou, Esther L. German, Jesús Reiné, Stephen B. Gordon, Simon P. Jochems, Jamie Rylance, Daniela M. Ferreira

## Abstract

*Streptococcus pneumoniae* (pneumococcus) is the most common identified bacterial cause of pneumonia, and the leading infectious cause of death in children under five years of age worldwide. Pneumococcal disease follows a seasonal pattern with increased incidence during winter. Pneumonia burden is also associated with poor air quality. Nasopharyngeal carriage of the bacterium is a pre-requisite of invasive disease.

We aimed to determine if susceptibility to nasopharyngeal pneumococcal carriage varied by season, and which environmental factors might explain such variation. We also evaluated the influence of sex on susceptibility of carriage. We collated data from five studies in which human volunteers underwent intranasal pneumococcal challenge. Generalised linear mixed effects models were used to identify factors associated with altered risk of carriage acquisition, specifically climate and air-quality data.

During 2011-2017, 374 healthy adults were challenged with type 6B pneumococcus. Odds of carriage were significantly lower in males (OR, 0.61; 95% CI, 0.40-0.92; *p* = 0.02), and higher with cooler temperatures (OR, 0.79; 95% CI, 0.63-0.99; p = 0.04). Likelihood of carriage also associated with lower concentrations of local fine particulate matter concentrations (PM_2.5_) and increased local rainfall.

In contrast to epidemiologic series, experimental challenge allowed us to test propensity to acquisition during controlled exposures; immunologic explanations for sex and climatic differences should be sought.

## Introduction

Infection with *Streptococcus pneumoniae* (pneumococcus) is characterised by initial colonisation in the nasopharynx. Nasopharyngeal carriage of pneumococcus is also the source of human-to-human transmission [1], with point prevalence estimated at 45-66% of infants and <10% of adults [1-3]. Most frequently, this is a transient state, although subsequent pathogenic infection can occur encompassing a spectrum of severity from self-resolving otitis media to life-threatening sepsis. The burden of pneumonia is particularly high; *S. pneumoniae* is the commonest identified bacterial pathogen. It is associated with a three-fold higher risk of mortality than non-pneumococcal pneumonia [4, 5], and is the major contributor to the annual 880,000 child pneumonia deaths [6].

Invasive pneumococcal disease is seasonal with peaks in winter most obvious in adult groups [7], and pneumococcal carriage rates in infants are significantly higher in cooler and drier months [8]. Around half of child pneumonia deaths have been attributed to poor air quality [6]. Short-term exposure to naturally occurring particulate matter (PM) has previously been associated with increased risk of hospital admission due to pneumonia, particularly in the elderly [9]. However, a more recent study involving univariate analysis showed that PM with a mean aerodynamic diameter ≤2.5μm (PM_2.5_), PM_1_ and PM_10_, along with relative humidity, temperature and solar radiation, were inversely correlated with rates of pneumococcal infection [10]. Conversely, SO_2_, NO_x_, NO_2_, NO and CO were found to be positively correlated with pneumococcal infection.

The experimental pneumococcal challenge programme in Liverpool (UK) provides data on the propensity to colonisation after instillation of *S. pneumoniae* into the nose. This offers a unique opportunity to study the seasonal and other factors which determine carriage given a known exposure timepoint and bacterial density. We used systematically collected data and participant demographics across several experimental studies to answer two main questions: Does the rate of susceptibility to nasopharyngeal pneumococcal carriage acquisition change throughout the year? Do environmental parameters affect carriage acquisition?

## Methods

A meta-analysis was conducted using data of five clinical trials carried out at the Clinical Research Unit, Liverpool School of Tropical Medicine between November 2011 and March 2017. All studies were approved by the sponsors and the North West NHS Research Ethics Committee (REC numbers: 11/NW/0592, 15/NW/0146, 14/NW/1460, 12/NW/0873). Inclusion criteria for all the studies were any person who spoke fluent English and was between the age of 18 and 50, except for 11/NW/0592, which had an upper age limit of 60. Exclusion criteria for the trials were contact with at risk individuals, current smoker or significant smoking history (>10 pack years), asthma or respiratory disease, chronic illness, pregnancy, penicillin allergy, involved in another clinical trial unless observational or in follow-up (non-interventional) phase and unable to give fully informed consent. None of the volunteers had received a pneumococcal vaccine. All volunteers provided written informed consent.

Participants were inoculated in each nostril with serotype 6B pneumococcus as previously described, with carriage defined as detection at any follow-up timepoint of serogroup 6 pneumococcus by classical microbiology with latex agglutination testing [11]. Follow-up sampling was performed over 1 month.

Contemporaneous air-quality data files from Speke, Liverpool were available from the Department for Environment Food & Rural Affairs. These data included measurements of air-quality determinants (PM_2.5_, PM_10_, volatile PM_10_, volatile PM_2.5_, non-volatile PM_2.5_, ozone, NO, NO_2_, NO as NO_2_ and SO_2_) which were similarly summarised as monthly averages for the period. Seasonality of climate and air quality data was confirmed (Supplementary Figures 1 and 2).

Regional climate data were obtained for 2011-2017 from the nearest Meteorological Office facility (located 69 miles West of Liverpool in Valley, Anglesey). Environmental data measurements were averaged for each climatic variable for every month of each year, including maximum and minimum temperatures (°C), rainfall (mm) and sun hours.

All data analysis and modelling were carried out using R software (R version 3.4.2). Studies were initially analysed using separately constructed generalised linear models. However, due to small sample sizes, data were pooled using a generalised linear mixed-effects (GLME) model, accounting for study effect, represented by the formula:

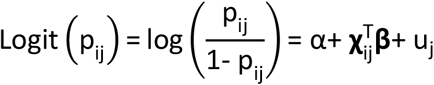

where α is a random intercept for each individual and u_j_ accounts for study effect it is reasonable to assume that *u* ∼ *Normal* (0,^2^).

Collinear variables were expected from climate and air-quality measures (Supplementary Figures 3 and 4); NO as NO_2_, volatile PM_2.5_, volatile PM_10_ and non-volatile PM_2.5_ were removed from the model to improve accuracy. Temperature measurements were represented in the model by the daily minimum. Where there was limited collinearity, but a strong prior evidence of association with carriage, variables were maintained in the model (PM_2.5_ and PM_10_, SO_2_ and NO and NO_2_). The GLME model included sex, age (categorical), inoculation month, minimum temperature, temperature difference, rainfall, sun hours, PM_10_, PM_2.5_, ozone, NO, NO_2_ and SO_2_. Where models did not converge, scaling was applied to covariates using the scale function in R [12]. Backwards elimination was used to remove non-informative variables and model fit was determined by minimising Akaike information criterion (AIC). Interactions between variables were assessed and covariance of model terms was determined using the ‘vcov’ function in the package ‘MASS’. Intraclass correlation (ICC) was generated using the ‘ICC’ function in the package ‘sjstats’.

## Results

### Study Cohort Demographics

Between November 2011 and March 2017, 407 volunteers were intranasally inoculated with pneumococcus serotype 6B across five clinical trials (Table 1, Figure 1). Natural carriers (carriage of non-serogroup 6 pneumococcus) were excluded from analysis (33/407 volunteers). Hence, a total of 374 volunteers were included in our analysis, 210 volunteers (56.2%) were female. Of these 374 volunteers, 171 (45.7%) became colonised. Most volunteers were recruited from university campuses in Liverpool and average volunteer age was 22 (87.7% of participants were aged 18-25). Most data were collected during winter months (October-March; 329/374) (Table 1).

**Table 1:**
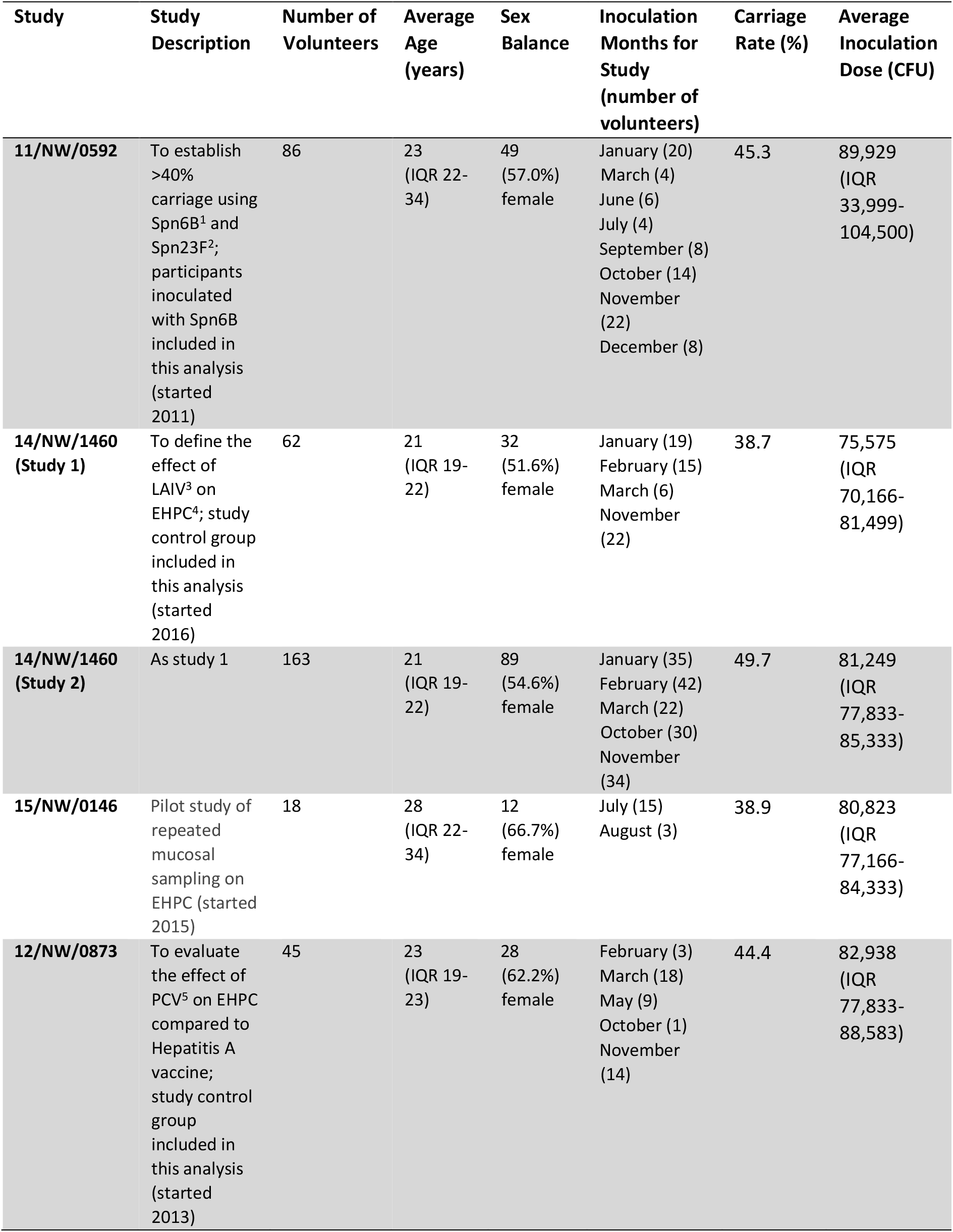
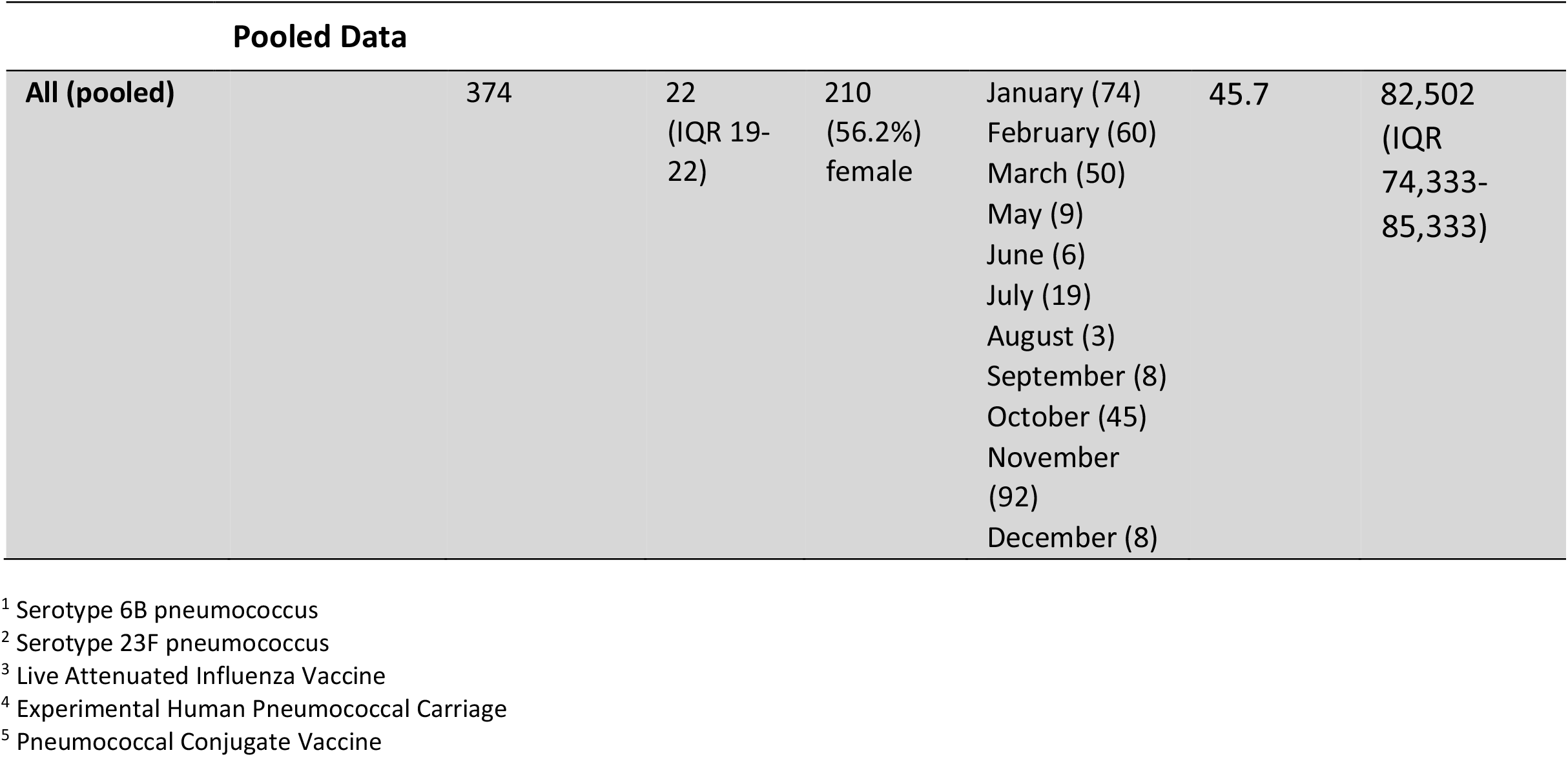
Demographics data for each clinical trial and pooled data.

**Figure 1:**
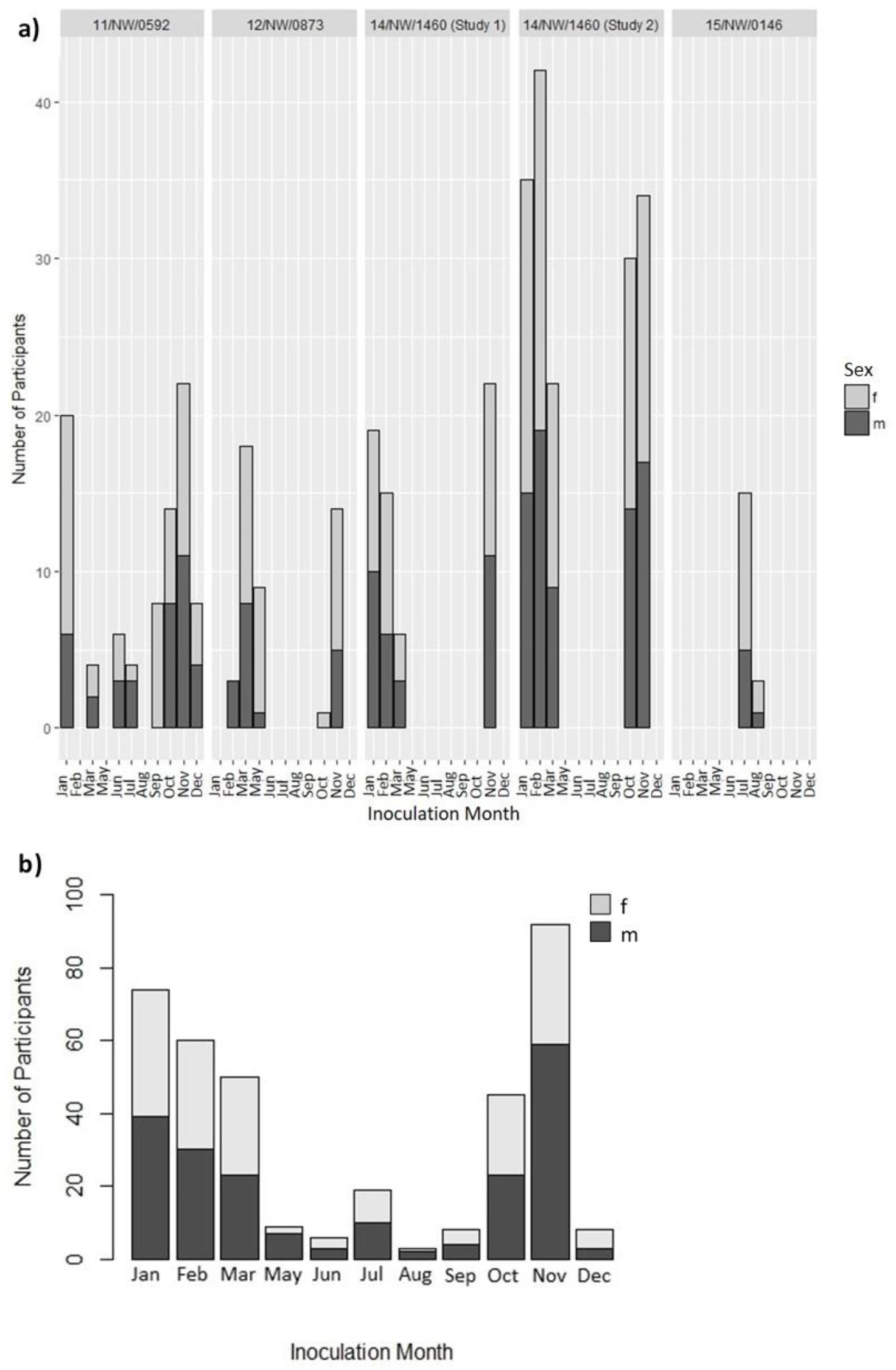
**a)** Bar plots showing distribution of data across months of the year for each study (m = male, grey, f = female, black). **b)** Distribution of data across months of the year for the pooled cohort (m = male, f = female). No volunteers were inoculated during the month of April in any study.

### Male sex associates with decreased susceptibility to pneumococcal carriage acquisition

First, we assessed whether time of the year was associated with carriage acquisition rates (Figure 2a). May, August and November had the lowest proportion of carriage of all months, below the overall average of 45.7% (22.2%, 33.3% and 35.9%, respectively) whilst December and March had the highest (62.5% and 54.0%, respectively). Carriage acquisition rates for remaining months ranged from 47.3-50.0%. Logistic regression analysis using a generalised linear model found no specific inoculation month was significantly associated with carriage acquisition. However, the sample sizes for the months April through September and December were small (Table 1, Figure 1, Figure 2a). As for inoculation month, age category did not significantly associate with carriage acquisition rates, although there was a lack of data for ages over 30 (Table 1, Figure 2b). Of the 210 female volunteers, 107 (51.0%) became colonised whereas, of the 164 male volunteers, 64 (39.0%) became colonised (Figures 2c and 2d). The number of males who became carriers following inoculation was significantly lower than the number of women who became colonised (Fisher’s exact test OR, 0.61; 95% CI, 0.41-0.93; *p* = 0.03).

**Figure 2:**
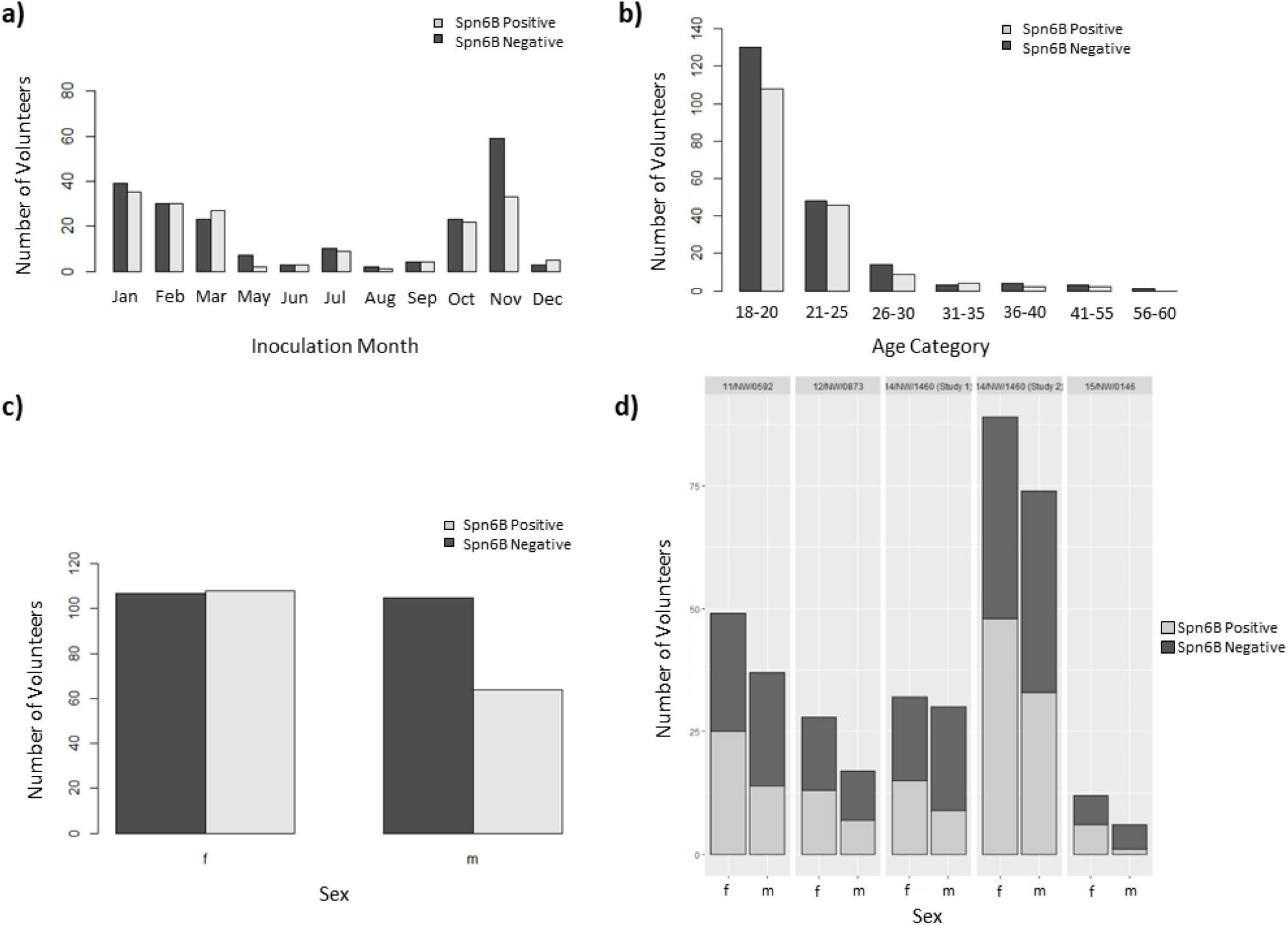
**a)** The number of carriage positive (grey) and carriage negative (black) volunteers in each inoculation month. **b)** The number of carriage positive and carriage negative volunteers in each age category. **c)** The number of female (f) and male (m) volunteers in the pooled dataset with positive (grey) and negative (black) pneumococcal carriage status. **d)** The number of female (f) and male (m) volunteers in each study with positive (grey) and negative (black) pneumococcal carriage status.

### Time taken to establish experimental carriage following nasal inoculation is not affected by season, age or sex

Notably, in 87.1% of the 171 volunteers who established carriage, colonisation was detected for the first time at the first sampling point following inoculation (day 2) (Figure 3) as compared to 10.5% at the second sampling point (day 6, 7 or 9) and 1.2% at the third and fourth sampling points (day 14 and day 27, respectively). Sex, age and inoculation month had no obvious effect on the day that carriage was first detected (Figure 3).

**Figure 3:**
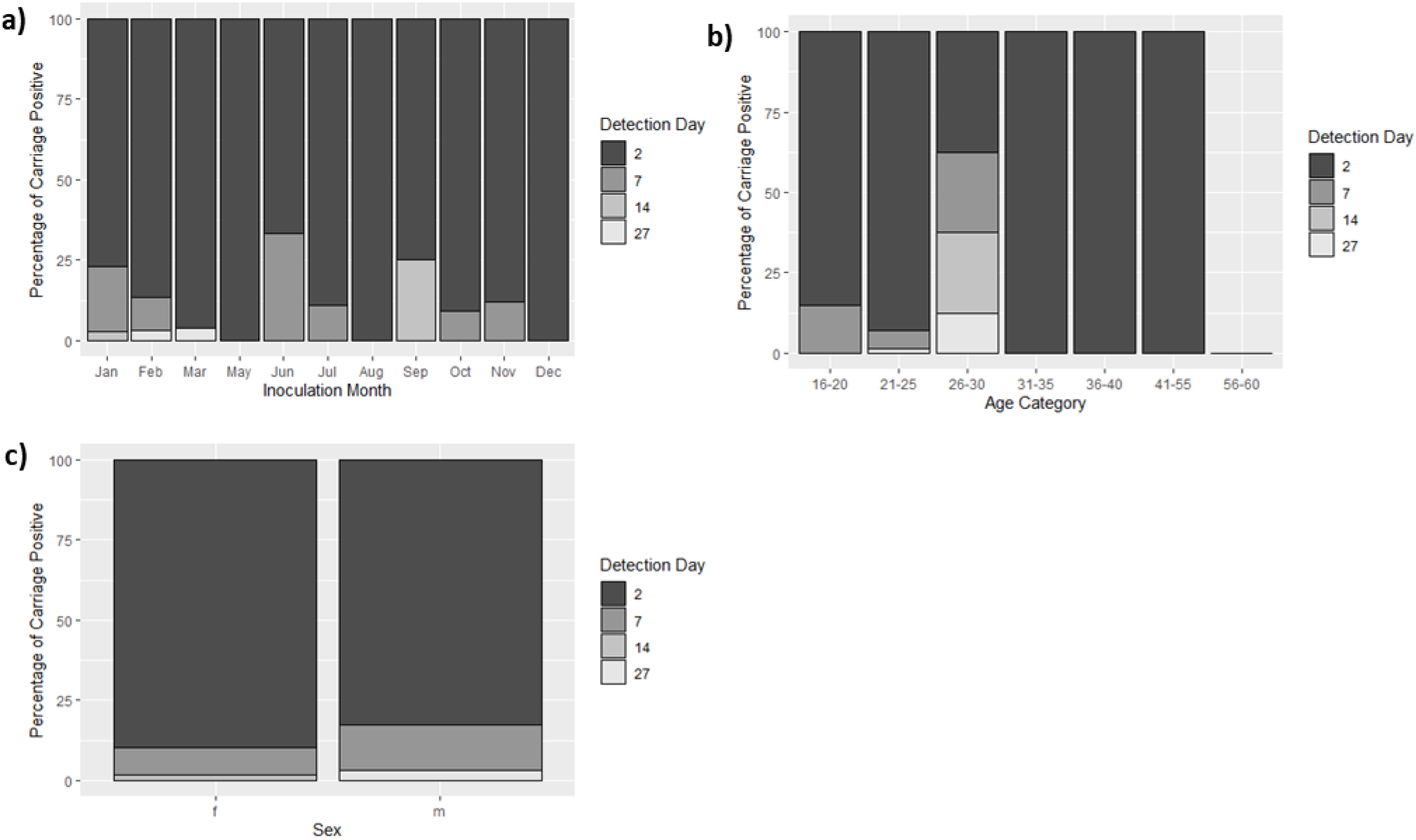
Percentage of carriage positive volunteers on which pneumococcal carriage was first detected 2 (black), 6, 7 or 9 (dark grey), 14 (light grey) or 27 (white) days after initial inoculation across each inoculation month **(a)**, each age category **(b)** and in females and males **(c)**.

### Statistical analysis and Generalised Linear Mixed Effects (GLME) model

To understand the impact of environmental variables, we then used a GLME model to assess the strength of association of age, gender, temperature, rainfall and air quality with carriage acquisition (Table 2).

**Table 2:** Log Odds, standard error values and *p*-values obtained from the generalised linear mixed-effects model.

|  | Logit<br>Coefficient<br>(Log Odds) | Standard<br>Error<br>(SE) | *p*-value |
| --- | --- | --- | --- |
| Sex (male) | -0.502 | 0.214 | 0.019 |
| PM<sub>2.5</sub><sup>1</sup> | -0.248 | 0.128 | 0.052 |
| Minimum<br>temperature | -0.239 | 0.118 | 0.043 |
| Rainfall | -0.175 | 0.113 | 0.124 |

<sup>1</sup> Particulate matter <2.5µm in diameter

Results from the GLME model found that males were significantly less likely to be colonised with pneumococcus than were females (OR, 0.61; 95% CI, 0.40-0.92; *p* = 0.02). Moreover, increased minimum temperature (OR, 0.79; 95% CI, 0.63-0.99; *p* = 0.04) was significantly associated with decreased likelihood of carriage acquisition rates. Although not significant, PM_2.5_ was also correlated with decreased odds of carriage acquisition (OR, 0.78; 95% CI, 0.61-1.00; *p* = 0.05), as was rainfall (OR, 0.84; 95% CI, 0.67-1.05; *p* = 0.12). The Akaike information criterion (AIC) of the model increased when these variables were removed, suggesting a potential link with colonisation.

As would be expected, there was collinearity between PM_2.5_, minimum temperature and rainfall (Table 3). Thus, the standard errors of parameter coefficient estimates are likely greater than they would be if the parameters were not correlated.

**Table 3:** Correlation matrix for variables included in GLME model.

|  | PM<sub>2.5</sub> | Minimum Temperature | Rainfall |
| --- | --- | --- | --- |
| PM<sub>2.5</sub><sup>1</sup> | 1 | 0.44 | 0.36 |
| Minimum Temperature | 0.44 | 1 | 0.11 |
| Rainfall | 0.36 | 0.11 | 1 |

The intraclass correlation (ICC) showed study grouping did not account for any residual variance in the model (ICC = 0.00%).

## Discussion

We associated subject sex and climatic variables with outcome following experimental human challenge studies conducted between 2011 and 2017, including 374 volunteers. The multivariate analysis conducted in this study found that male sex is significantly associated with reduced odds of establishing pneumococcal carriage. Increases in minimum temperature were significantly associated with decreased rates of colonisation, thus odds of colonisation were significantly increased with cooler temperatures, concurring with findings of previous studies [7, 13, 14]. Increased PM_2.5_ was inversely correlated with pneumococcal carriage acquisition, although this was not significant. Similarly, although not significantly, odds of colonisation were found to be at least weakly inversely correlated with increased rainfall, corresponding to previous results [8]. Additional environmental variables for which data were obtained were not found to improve model fit. Nor did the inclusion of age category. The positively skewed age distribution limited our power to detect differences in pneumococcal carriage acquisition between the younger and older volunteers. Across all the clinical trials included in this analysis, volunteer cohorts were predominantly university students, which explains the paucity of data for volunteers over 30 years of age and for months when students are away from university (April through September for Easter and Summer holidays and December for Winter holiday).

The finding in our study that males had significantly lower likelihood of pneumococcal carriage acquisition than females is supported by previous studies showing that females are more susceptible than males to infections of the upper respiratory tract whereas, males are more susceptible to lower respiratory tract infections [15]. Nevertheless, the results of this study differ from previous analyses, which have found that male sex is significantly associated with increased odds of nasopharyngeal pneumococcal carriage [16, 17] or that there is no significant difference in the prevalence of carriage between sexes [13]. A potential explanation for this discrepancy is that previous studies analysing sex-linked odds of carriage have included participants under the age of 18, whereas our study included adults over 18 years of age only. It is unlikely that there are differences in the sex-linked odds of carriage in children, whereas adults and adolescents will exhibit hormone effects on the immune system. These have been recently reviewed in the lung, with androgens generally considered anti-inflammatory and oestrogens pro-inflammatory [18] in the context of chronic lung disease. The delicate balance that determines mucosal immunity in response to acute challenge will differ from that in chronic inflammation and is a focus of our group’s work [19]. Furthermore, our study uses a controlled model of infection, with important variables including geographical location controlled. Previous studies considered natural carriage of various serotypes within various populations in countries such as Brazil and sub-Saharan Africa where social factors have different effects on interaction, contact and social crowding. Using a controlled model of infection thereby enables us to determine propensity of colonisation based on sex while controlling for other variables which may interfere with this finding.

We observed that increased PM_2.5_ concentration associated with a decreased likelihood of carriage acquisition. This is not counter-intuitive and has been shown in a mouse model [20]. The mucosal defence against pneumococcus is dependent on neutrophil activation and smoke potentially activates neutrophils in the mucosa. It has been well established that fine and ultrafine particulates are detrimental to lung and cardiovascular health, both in the long and short-term, increasing morbidity and mortality [21-24] by a cytokine based pro-inflammatory mechanism. The burden of pneumococcal infection, particularly in the elderly, is worsened following exposure to ambient PM_2.5_ and exposure to PM may increase pneumonia-related mortality by supressing immune functions such as macrophage responsiveness [9, 25]. Coarse particulates (>8μm) have greater impact on the upper respiratory tract, depositing in the pharynx, larynx and trachea in a size-dependent manner, whilst fine particulates (<1–3μm) penetrate lung tissue and deposit in the alveoli [26, 27]. When air pollution particles interact with alveolar macrophages, phagocytosis, release of inflammatory mediators and oxidant production is triggered [28]. However, alveolar macrophage function, particularly phagocytosis of pneumococci and mycobacteria, is impaired by increased exposure to particulates [19]. Interestingly, although internalisation and phagocytosis decreases, alveolar macrophage binding to pneumococcus has been shown to be enhanced by PM_2.5_ [28]. The effect of exposure to PM on the nasal mucosa has been less well documented, although some murine and human studies have demonstrated that exposure to particulates induces recruitment of immune cells to the nasal mucosa, promoting inflammation [29-32]. Thus, it is conceivable that increased inflammation related to higher levels of PM_2.5_ led to a transient state of protection against carriage acquisition.

Notably, preliminary data has shown that increased exposure to air pollution promotes expression of PAF receptors in the nasal epithelium, and thus it might be expected that this would enhance pneumococcal binding to the nasal epithelium and subsequent carriage [33]. On the other hand, increased exposure of the nasal epithelium to PM_2.5_ stimulates increased IL-8 production [34]. IL-8 is a pro-inflammatory chemoattractant and promotes the recruitment of neutrophils to the nasal mucosa, which play an important role in pneumococcal clearance [35]. Thus, it could be postulated that in this way pneumococcal carriage is inhibited.

Mice models of nasopharyngeal carriage of pneumococcus have exhibited significantly increased pneumococcal density in the nasopharynx following exposure to dust (representative of particulates <10μm) compared to normal bacterial colonised control mice. In addition, exposure to dust triggered local inflammation in the nasopharynx and promoted a more invasive pneumococcal phenotype [36]. Dust exposure also resulted in significantly reduced phagocyte-mediated bacterial killing. Similarly, a 2017 study showed that in murine models of nasopharyngeal colonisation, black carbon, a major component of PM, promoted the spread of pneumococcus from the nasopharynx to the lungs [37]. However, the same degree of colonisation was observed in the upper respiratory tracts of mice exposed to both black carbon and pneumococcus and those infected with pneumococcus only. Interestingly, black carbon was found to induce a biological effect on non-capsulated pneumococcus resulting in a change in biofilm structure. In contrast, our study found that PM_2.5_ elicited a protective effect against nasopharyngeal pneumococcal colonisation. Markedly, ours is the first study to model the effects of ambient PM on experimental pneumococcal carriage in humans. Similarly, previous univariate analysis found that PM was associated with reduced rates of pneumococcal infection [10]. Whilst no definitive explanation may be offered at this point, as to why PM_2.5_ might impair colonisation, it could be hypothesised that the presence of particulates might prevent bacteria from establishing colonisation due to the enhanced presence of pro-inflammatory cells in the nasopharynx. Furthermore, it is important to consider that that the chemical composition of PM could alter its biological effects and thus location is an important factor [21, 38-40].

Although measures were taken to limit the inclusion of collinear variables in the model, this does not allow dissection of the independent effects. Unknown confounders, for example concomitant respiratory viral infections, are likely to play are part in these complex relationships. The scope of work was limited by the range of data elements incorporated into previous studies. While we excluded smokers from the study, for example, we have no information on exposure to second-hand smoke. Our analysis at aggregate environmental data was also limited in resolution. Nevertheless, volunteer cohorts were relatively homogeneous by occupation, general health status, non-smoking status, and inoculum dose which gives confidence in the internal consistency of our findings.

In conclusion, this study found that males are significantly less likely than females to be colonised in the nasopharynx following experimental challenge with serotype 6B pneumococcus. Furthermore, nasopharyngeal pneumococcal carriage acquisition was found to be positively correlated with cooler temperatures, decreased levels of PM_2.5_ in the environment and higher rainfall. Further studies and analysis are needed to better ascertain correlation of pneumococcal acquisition with environmental factors in order to establish how the environment, particularly air quality, could affect likelihood of pneumococcal carriage acquisition in the community. Further work is being conducted to establish how sex might alter immunological responses to pneumococcus.

## Data Availability

Data from trials mentioned in the manuscript have been published prevously. For this analysis, we used collated colonisation data from our challenge model trials. Air-quality data files from Speke, Liverpool were available from the Department for Environment Food & Rural Affairs. Regional climate data were obtained for 2011-2017 from the nearest Meteorological Office facility (located 69 miles West of Liverpool in Valley, Anglesey).

## Acknowledgements

KSC is funded by the MRC Doctoral Training Partnership. DMF, AMC, SBG, SPJ, JR, CS, BU and KSC are members of the Human Infection Challenge Network for Vaccine Development (HIC-Vac), which is funded by the GCRF Networks in Vaccines Research and Development, which was co-funded by the MRC and BBSRC. The challenge studies were funded by the MRC (grant MR/M011569/1) awarded to SBG and DMF and the Bill and Melinda Gates Foundation (grant OPP1117728) awarded to DMF. We would like to acknowledge Dr Helen Hill, Sister Angela Hyder-Wright, Dr Jenna Gritzfeld, Dr Hugh Adler, Dr Seher Zaidi and Dr Victoria Connor for their contribution to the EHPC trials referred to in this work.

---

**Supplementary Figure 1:**
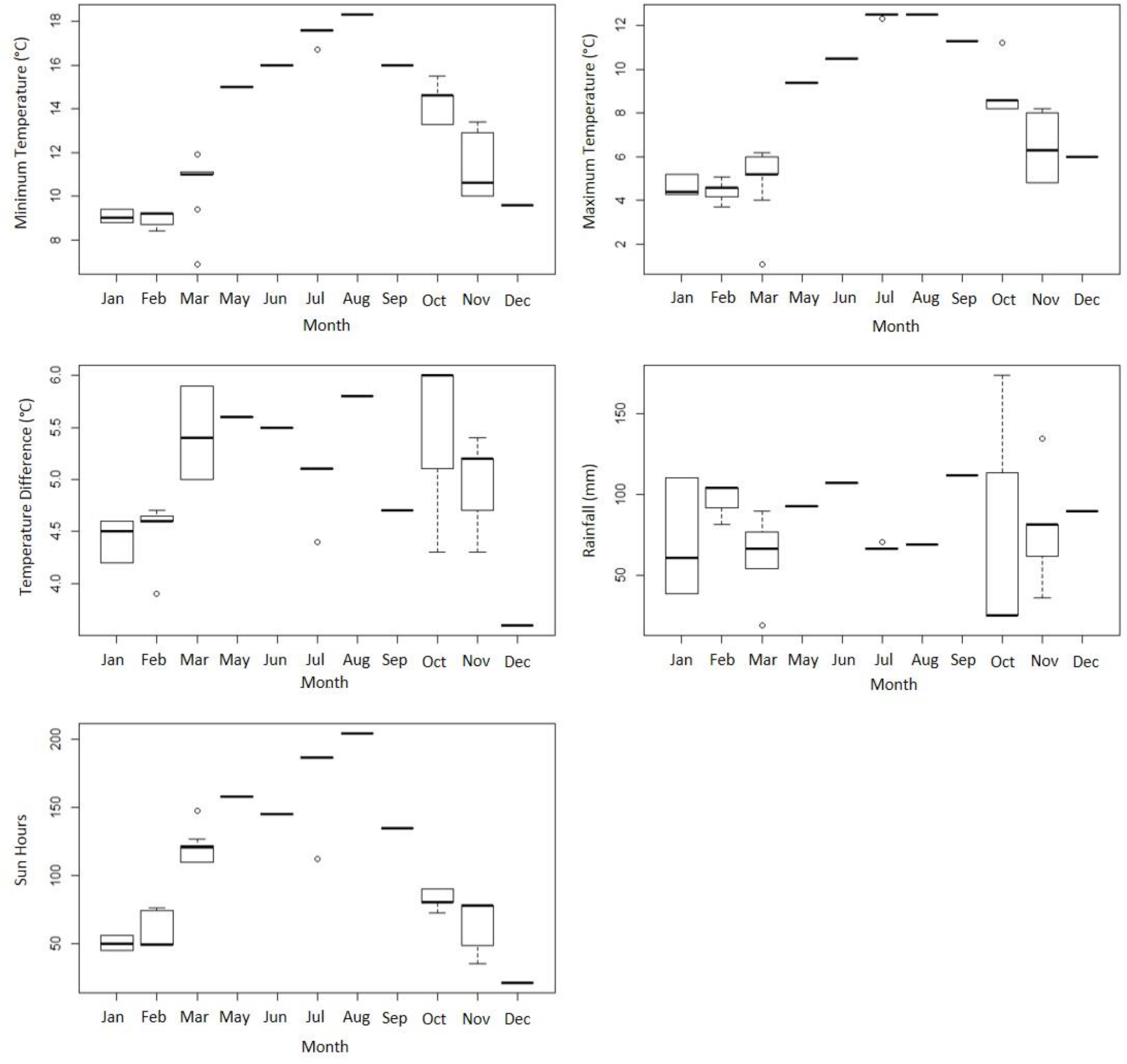
Aggregated monthly average minimum temperature (°C), maximum temperature (°C), temperature difference (°C), rainfall (mm) and sun hours from November 2011 to March 2017 (excluding month of April). Data obtained from the Met Office Valley, Anglesey monitoring site.

**Supplementary Figure 2:**
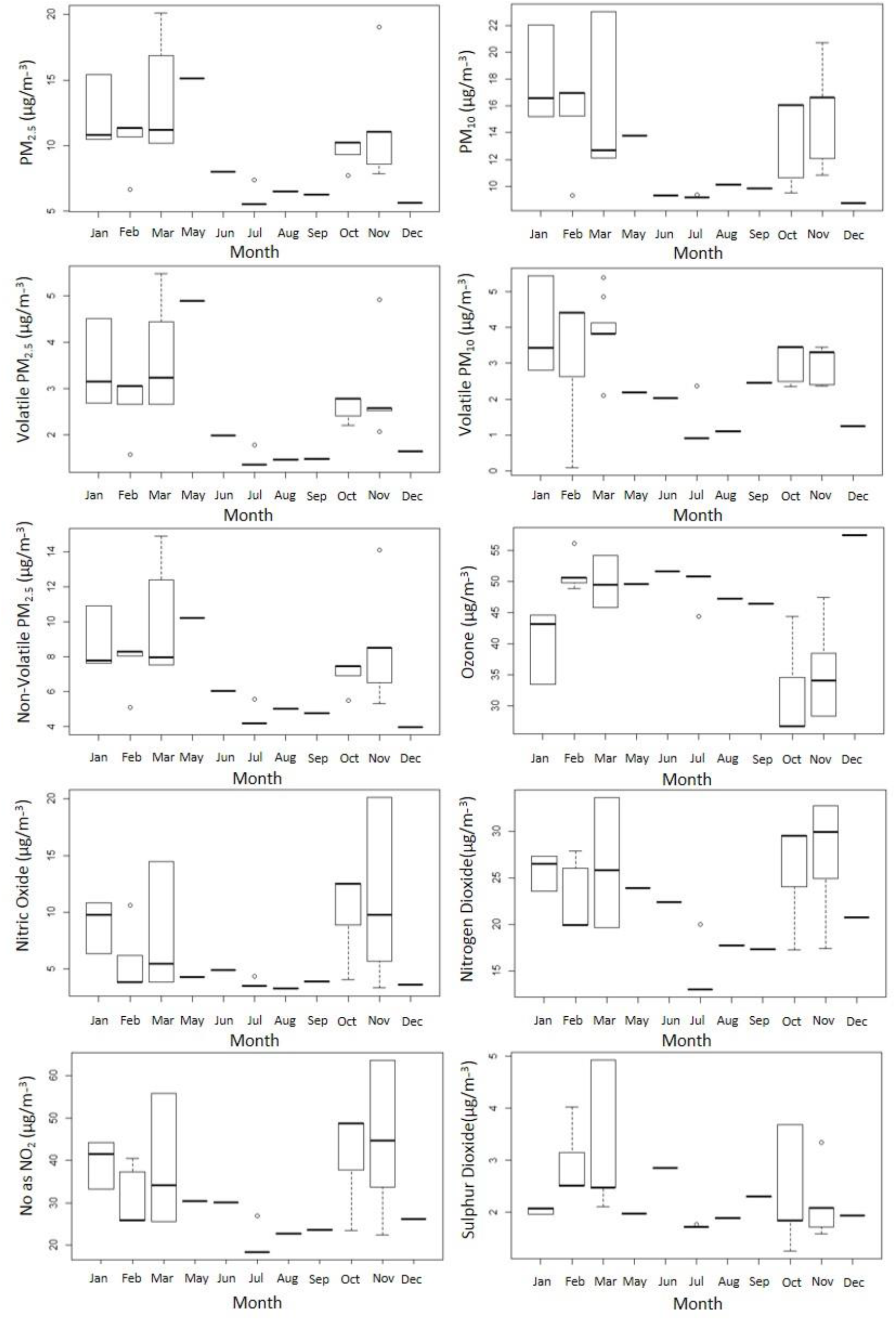
Aggregated monthly average (µg/m^-3^) PM_2.5_ and PM_10_, volatile and non-volatile PM_2.5_, volatile PM_10_, ozone, NO, NO_2_, NO as NO_2_ and SO_2_ from November 2011 to March 2017 (excluding month of April). Data obtained from DEFRA Speke, Liverpool monitoring site.

**Supplementary Figure 3:**
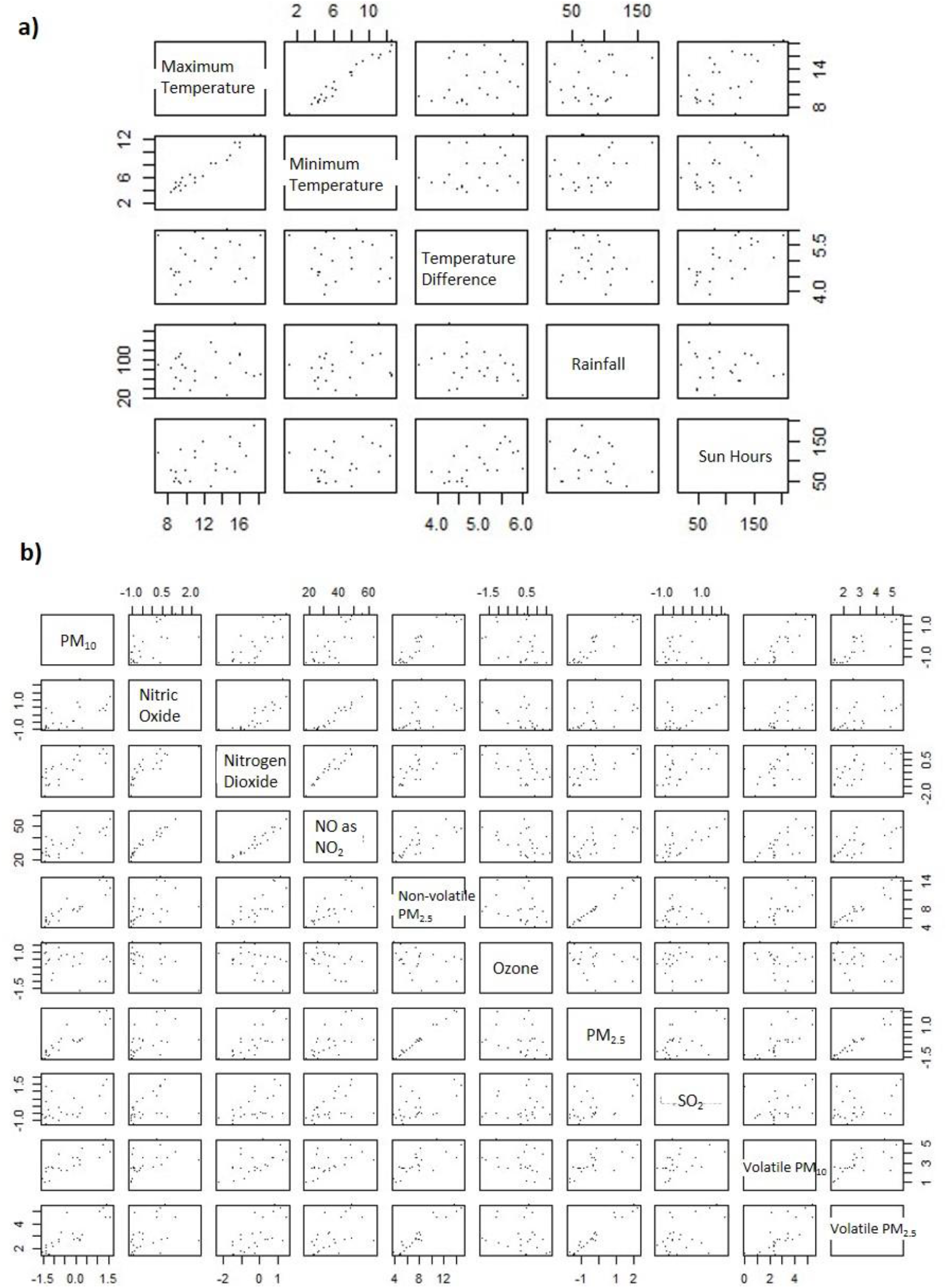
**a)** Plots to determine collinearity of climate variables maximum temperature (°C), minimum temperature (°C), temperature difference (°C), rainfall (mm) and sun hours. Maximum and minimum temperature were strongly correlated. **b)** Plots to determine collinearity of air-quality variables (µg/m^-3^) PM_10_, NO, NO_2_, NO as NO_2_, non-volatile PM_2.5_, ozone, PM_2.5_, SO_2_, volatile PM_10_ and volatile PM_2.5_. NO as NO_2_ data mimics that of NO and NO_2_ separately. Non-volatile PM_2.5_ and volatile PM_2.5_ data were found to be combined within total PM_2.5_ data. Similarly, PM_10_ data was strongly correlated with volatile PM_10_ data. Limited collinearity exists between PM_2.5_ and PM_10_ and between SO_2_, NO and NO_2_.

**Supplementary Figure 4:**
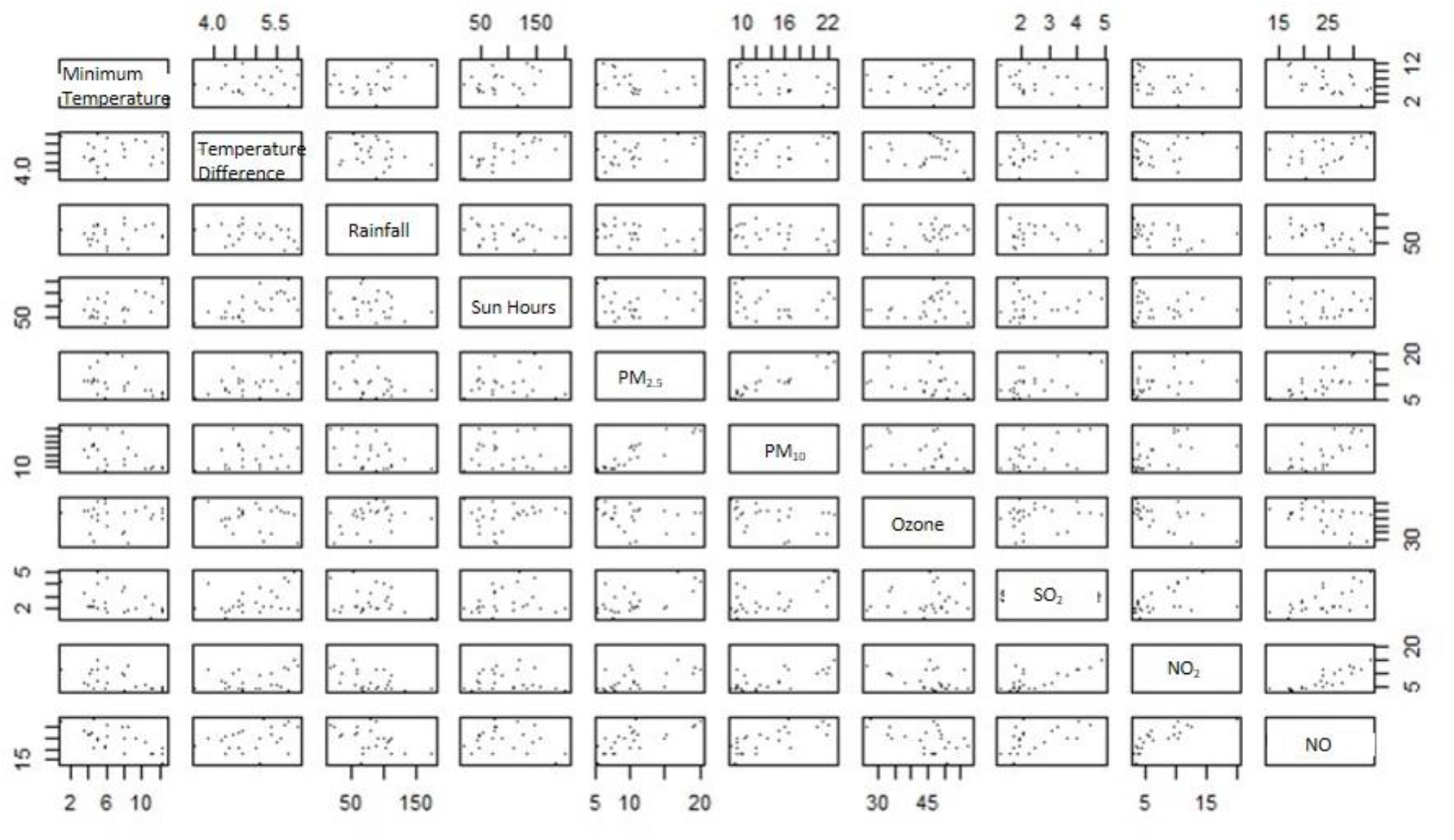
Plots to determine collinearity of variables minimum temperature (°C), temperature difference (°C), rainfall (mm), sun hours, PM_2.5_ (µg/m^-3^), PM_10_ (µg/m^-3^), ozone (µg/m^-3^), SO_2_ (µg/m^-3^), NO (µg/m^-3^) and NO_2_ (µg/m^-3^).

